# GWAS-based Machine Learning for Prediction of Age-Related Macular Degeneration Risk

**DOI:** 10.1101/19006155

**Authors:** Qi Yan, Yale Jiang, Heng Huang, Anand Swaroop, Emily Y. Chew, Daniel E. Weeks, Wei Chen, Ying Ding

**Affiliations:** Division of Pulmonary Medicine, Allergy and Immunology, Department of Pediatrics, Children’s Hospital of Pittsburgh of UPMC, University of Pittsburgh, Pittsburgh, PA; School of Medicine, Tsinghua University, Beijing, China; Department of Electrical and Computer Engineering, Swanson School of Engineering, University of Pittsburgh, PA; Department of Biomedical Informatics, School of Medicine, University of Pittsburgh, PA; Neurobiology Neurodegeneration and Repair Laboratory, National Eye Institute, National Institutes of Health, Bethesda, MD; Division of Epidemiology and Clinical Applications, National Eye Institute, National Institutes of Health, Bethesda, MD; Department of Human Genetics, Graduate School of Public Health, University of Pittsburgh, PA; Department of Biostatistics, Graduate School of Public Health, University of Pittsburgh, Pittsburgh, PA

## Abstract

Numerous independent susceptibility variants have been identified for Age-related macular degeneration (AMD) by genome-wide association studies (GWAS). Since advanced AMD is currently incurable, an accurate prediction of a person’s AMD risk using genetic information is desirable for early diagnosis and clinical management. In this study, genotype data of 32,215 Caucasian individuals with age above 50 years from the International AMD Genomics Consortium in dbGAP were used to establish and validate prediction models for AMD risk using four different machine learning approaches: neural network, lasso regression, support vector machine, and random forest. A standard logistic regression model was also considered using a genetic risk score. To identify feature SNPs for AMD prediction models, we selected the genome-wide significant SNPs from GWAS. All methods achieved good performance for predicting normal controls versus advanced AMD cases (AUC=0.81∼0.82 in a separate test dataset) and normal controls versus any AMD (AUC=0.78∼0.79). By applying the state-of-art machine learning approaches on the large AMD GWAS data, the predictive models we established can provide an accurate estimation of an individual’s AMD risk profile across the person’s lifespan based on a comprehensive genetic information.

## INTRODUCTION

Age-related macular degeneration (AMD) is a multifactorial neurodegenerative disease, which is a leading cause of vision loss among the elderly in the developed countries.^1; 2^ The disease affects the central vision and is progressive, starting with the appearance of drusen (i.e., the yellow or white deposits in the eye) and eventually leading to advanced AMD forms: wet AMD (choroidal neovascularization [CNV]) and dry AMD (geographic atrophy [GA]) ^3^. Patients can progress to one or both forms of advanced AMD. Some patients with early AMD maintain good vision for a long time without progressing to advanced AMD, while others quickly developed advanced AMD.

In 2005, Fisher et al reported that the *CFH* gene on chromosome 1 and *ARMS2/HTRA1* genes on chromosome 10 were the most replicated gene regions associated with AMD ^4^. Later, with the advances of technology, multiple genome-wide association studies (GWAS) were conducted to examine the association between AMD and a genome-wide set of single nucleotide polymorphisms (SNPs). In 2016, the International AMD Genomics Consortium identified or confirmed a total of 34 loci with 52 independent genetic variants to be associated with advanced AMD risk ^5^. From this study, the phenotype and genotypes of 35,358 subjects were uploaded to dbGaP (phs001039.v1.p1) and the majority of them are Caucasians. Multiple studies demonstrated that the same AMD susceptibility loci were more strongly associated with AMD in Caucasians than in other ethnic groups ^6-8^.

Since advanced AMD is currently incurable, an accurate prediction of a person’s risk for (advanced) AMD using genetic information at a young age is desirable for early diagnosis and clinical management. In this study, our objective is to establish and validate prediction models for AMD risk based on genetic variants given any future age of a subject using the largest publicly available data from dbGaP (phs001039.v1.p1).

## METHODS

### Sample description and genotype data

The study subjects are from the International Age-Related Macular Degeneration Genomics Consortium - Exome Chip Experiment dbGaP data set (phs001039.v1.p1), which gathered samples from 26 studies. There are 32,215 Caucasians over 50 years of the total 35,358 subjects. Specifically, of the Caucasian subjects, 14,348 are normal controls, 5,290 are intermediate AMD cases, 2,644 are GA cases, 8,430 are CNV cases and 1,503 are GA/CNV mixed cases. Genotypes were imputed with the 1000 Genomes Project as the reference panel. A total of 13,503,037 genetic variants are included. The detailed subject recruitment, ascertainment of AMD severity and genotyping procedures were reported elsewhere ^5^.

In addition, we extracted a set of 383 Caucasian subjects (mean age = 62.30 ± 5.61) with macular degeneration (i.e., all cases) and above 50 years from UK Biobank^9^ as an independent test dataset. UK Biobank is the largest and most complete European Biobank available at present. All people in UK Biobank were aged from 40 to 69 years, which were much younger than the dbGaP AMD subjects (mean age = 76.32 ± 8.24). This may indicate that the AMD cohort from the UK Biobank only represents a narrow age range in Caucasians.

### Scenarios

We considered two main classification scenarios: 1. normal controls versus advanced AMD cases; and 2. normal controls versus any AMD cases (i.e., both intermediate and advanced AMD). In addition, we also considered another two binary outcome classification scenarios in the supplementary material: 3. normal controls versus intermediate AMD cases; and 4. intermediate AMD cases versus advanced AMD cases.

### Feature SNPs selection

For each of the four aforementioned classification scenarios, the data were randomly divided into a test dataset of 5,000 samples and a training dataset of the remaining samples. The training and test datasets were the same for all methods. We used the training dataset only to conduct the GWAS analysis and further used *p*-value to select feature SNPs as inputs for prediction models. The test dataset remains intact, and was saved for the final prediction performance evaluation. For all the classification scenarios, the GWAS was conducted using a logistic regression under an additive genetic model, adjusting for age, gender and the first two principal components calculated based on genotypes (for controlling population stratification). We then selected genome-wide significant SNPs with *P* < 5×10^−8^ as the feature SNPs in two ways. In the primary list of feature SNPs, only the top one SNPs from each of the significant loci were selected. In a secondary list of feature SNPs, all genome-wide significant SNPs were selected. In another secondary SNP list, all SNPs with *P* < 1×10^−5^ were selected. We only considered SNPs with minor allele frequency (MAF) greater than 0.01. In addition to the selected SNPs, we also included age as a predictor as it is known to be associated with AMD risk.

### Machine learning methods

We considered four machine learning methods in this study: neural network (NN), lasso regression (Lasso), support vector machine (SVM), and random forest (RF). As a comparison, we also fitted a standard logistic regression using a genetic risk score (GRS).

First, a multi-layer feedforward neural network was implemented using *Keras* ^10^. All layers were fully connected (Figure S1). We used two hidden layers with 16 nodes each and L1 norm regularization with lambda = 0.0001 after tuning multiple times at the input layer. Since NN can learn the complex relationship between predictors and outcomes, it was expected to be superior to the ones based on a linear relationship (such as the standard logistic regression with a lasso penalty). NN is often considered as a “black box” due to its complex inner architecture. To better interpret the predictions, LIME (Local Interpretable Model-Agnostic Explanations) was applied, which attempts to understand the model by perturbing the input of data samples and understanding how the predictions change. A 10-fold cross-validation was performed within the training dataset to find the best epoch (i.e., iteration) with the lowest loss for evaluation using the test dataset. Second, a lasso regression was implemented using the R function *glmnet* with the same value of the tuning parameter lambda ^11^. Note that the lambda values in both NN and Lasso were the tuning parameters for the L1 norm penalty. They could be different values. Since different lambda values led to similar results, for the sake of simplicity, we used the same lambda value for NN and Lasso. Moreover, linear SVM and RF were implemented using the R package *caret* ^12^. In addition, we computed a genetic risk score, 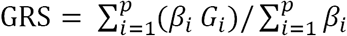, where *β*_*i*_ is the log(Odds Ratio) of the risk variant *i* as claculated from our GWAS results (similar approach was described in Ding *et al*. ^13^) and *G*_*i*_ is the corresponding genotype (coded as 0, 1 and 2: copies of risk allele). Here the *p* number of variants in GRS were the genome-wide significant SNPs (one from each independent locus) and this same set of SNPs were used in other machine learning approaches were used to calculate the GRS. Note that in this coding all *β*_*i*_ are positive and GRS ranges from 0 to 2. Then, a standard logistic regression was fitted with the GRS and age as the predictors. We refer to this method as GRS. For the binary outcome classification, we calculated the AUC (area under the curve) of Receiver Operator Characteristic (ROC) curves as the primary performance metric. Additionally the Brier score ^14^ was used as another metric, where the lower the Brier score is the better the model predicts. The useful benchmark value for the Brier score is 33%, which corresponds to predicting the risk by a random number drawn from a Uniform[0, 1] distribution. Model performance was evaluated in the separate test datasets.

## RESULTS

### Study data characteristics

The detailed demographic and clinical characteristics of the participants were described elsewhere ^5^. In this study, we used Caucasian participants with an age greater than 50 years old. The total sample size was 32,215, the mean age was 73.8 (SD = 9.3) years, and women comprised 57.6% (N = 18,554) of the cohort (Table 1). There were 14,348 normal controls, 5,290 intermediate AMD cases and 12,577 advanced AMD cases. As the AMD severity increased from no to intermediate to advanced AMD the mean age in those groups increased from 70.6±9.5 to 74.7±8.5 to 77.0±8.0 years (Table 1 and Figure S2). The percentage of women among the intermediate and advanced AMD cases (59.2% and 58.9%) was higher than observed in the normal controls (55.9%, Table 1).

**Table 1.** Characteristics summary

|  | N | Females, N (%) | Age (Mean±SD) |
| --- | --- | --- | --- |
| All | 32,215 | 18,554 (57.6%) | 73.8±9.3 |
| Normal controls | 14,348 | 8,021 (55.9%) | 70.6±9.5 |
| Intermediate AMD cases | 5,290 | 3,131 (59.2%) | 74.7±8.5 |
| Advanced AMD cases | 12,577 | 7,402 (58.9%) | 77.0±8.0 |

### Feature SNPs selection from GWAS of AMD

As shown in Figures 1 and S3, and Table S1, the Scenario 1 GWAS of normal controls versus advanced AMD cases resulted in the most genome-wide significant (*P* < 5×10^−8^) loci (18 loci [*CFH, ADAMTS9-AS, COL8A1, CFI, C9, C2/CFB/SKIV2L, VEGFA, ARMS2/HTRA1, ACAD10, B3GALTL, LIPC, CETP, CTRB2/CTRB1, C3, APOE, C20orf85, SYN3/TIMP3, SLC16A8*] that include 5,233 SNPs). All these loci were reported in Fritsch *et al*. ^5^, which also compared normal controls and advanced AMD cases. We did not capture all of the Fritsch *et al*. ^5^ previously reported loci, because we only used the training set in our GWAS in order to keep the test set untouched, and only used common variants with MAF > 0.01. The Scenario 2 GWAS of normal controls versus any AMD cases also showed many significant loci (16 loci with 5,553 SNPs) and most of them were in the Scenario 1 GWAS too. However, *TNFRSF10A* from chromosome 8 and *SMG6* from chromosome 17 were newly identified, which were not reported in Fritsch *et al*. ^5^, possibly because of the addition of intermediate AMD cases to the advanced cases. The Scenario 3 GWAS of normal controls versus intermediate AMD cases (4 loci with 1,583 SNPs) and the Scenario 4 GWAS of intermediate AMD cases versus advanced AMD cases (5 loci with 1,228 SNPs) identified fewer significant loci, as the intermediate AMD category typically contains individuals with a wide range of disease severity, which could be close to either no or advanced AMD. The power could be another issue due to a much smaller sample size of intermediate AMD cases. Although few loci were detected, the Scenario 4 GWAS identified *ABHD2* from chromosome 15, which was not reported in Fritsch *et al*. ^5^ This gene could be useful for differentiating intermediate and advanced AMD. In each scenario, we used the corresponding significant SNPs in the prediction model.

**Figure 1.**
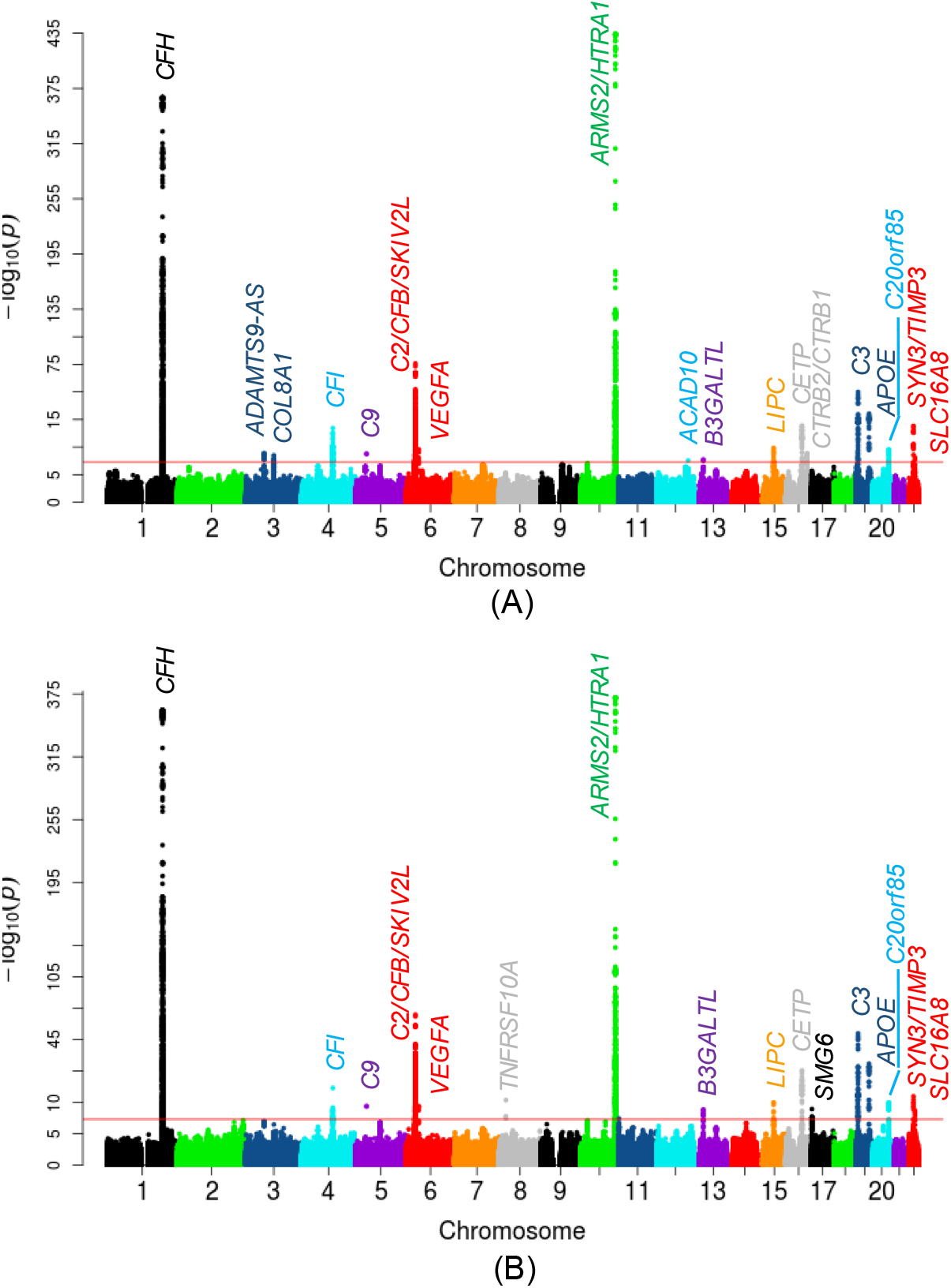
Manhattan plots of GWAS results. (A) Scenario 1: normal controls vs. advanced AMD cases, and (B) Scenario 2: normal controls vs. any AMD cases. The red horizontal line indicates the significance threshold (*P* = 5 × 10^−8^).

### Prediction performance

In our primary list of feature SNPs, we used the top one SNP from each of the genome-wide significant loci and age as predictors. Five model approaches including NN, Lasso, SVM, RF and GRS were conducted for each scenario. Each model was trained in the training set and evaluated in the test set. The AUC values and Brier scores based on the test set are presented in Table 2. The ROC curves and 95% CI of the AUC using the DeLong method ^15^ were also reported in Figures 2 and S4. Scenario 1 showed overall good predictions (AUCs between 0.81 and 0.82 for all five approaches). For Scenarios 3 and 4, all five prediction methods did not perform well (AUCs between 0.61 and 0.68). The reasons could be the fact that a wide range of samples fell into the category of intermediate AMD, which could be close to either controls or advanced AMD cases. Scenarios 2 also showed reasonably good performance (AUCs = 0.78). The density curves of predicted risks were generated and shown in Figures 3 and S5. Such plots allow us to visually examine the two counterparts from each comparison scenario separately. Similar to the AUC results, the Scenarios 1 and 2 showed clear separation. On the contrary, Scenarios 3 and 4 led to ambiguous results. The feature importance heatmaps from LIME (Figures 4 and S6) for NN further indicated that *CFH* and *AMRS2/HTRA1* contributed the most to the predictions (marked with darker colors). Note that the green vertical lines indicate that the feature supports the predicted classification for that subject and the red vertical lines indicate that the feature contradicts the predicted classification. In addition, all SNPs are in the same scale (additive model: 0 ∼ 2), but age is in a different scale (> 50 years). Thus, although the color of age looks light, it is a strong predictor. We also investigated the age effect on AMD risk by predicting a test dataset with age from 50 to 90 and all SNPs with common homozygous genotypes. The results (Figure S7) showed that both advanced and any AMD risks increased as age increased.

**Table 2.** AUC values (95% CI) and Brier scores (95% CI) of the prediction of Scenario 1. normal controls vs. advanced AMD cases, and Scenario 2. normal controls vs. any AMD cases

|  | normal controls vs. advanced AMD cases |  | normal controls vs. any AMD cases |  |
| --- | --- | --- | --- | --- |
|  | AUC | Brier score | AUC | Brier score |
| NN | 0.82 (0.81-0.83) | 0.17 (0.17-0.18) | 0.78 (0.77-0.80) | 0.19 (0.19-0.20) |
| Lasso | 0.82 (0.81-0.83) | 0.17 (0.17-0.18) | 0.78 (0.77-0.80) | 0.19 (0.18-0.19) |
| SVM | 0.82 (0.81-0.83) | 0.17 (0.17-0.18) | 0.78 (0.77-0.80) | 0.19 (0.18-0.19) |
| RF | 0.81 (0.80-0.82) | 0.18 (0.17-0.18) | 0.78 (0.76-0.79) | 0.19 (0.19-0.20) |
| GRS | 0.82 (0.81-0.83) | 0.17 (0.17-0.18) | 0.78 (0.77-0.80) | 0.19 (0.18-0.19) |

AUC 95% CI uses the DeLong method <sup>15</sup>; Brier score 95% CI uses bootstrap method.

**Figure 2.**
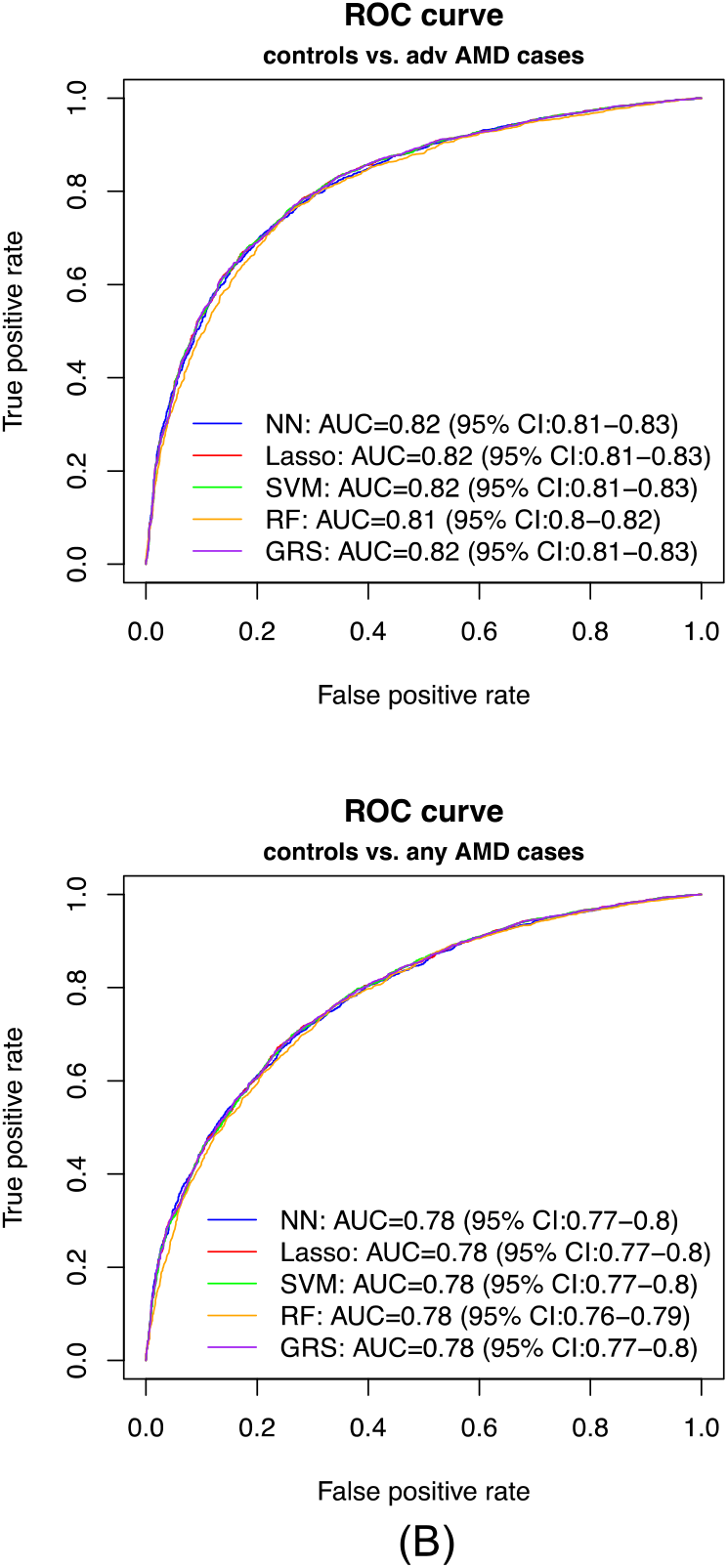
ROC curves of the predicted risk. (A) Scenario 1: normal controls vs. advanced AMD cases, and (B) Scenario 2: normal controls vs. any AMD cases.

**Figure 3.**
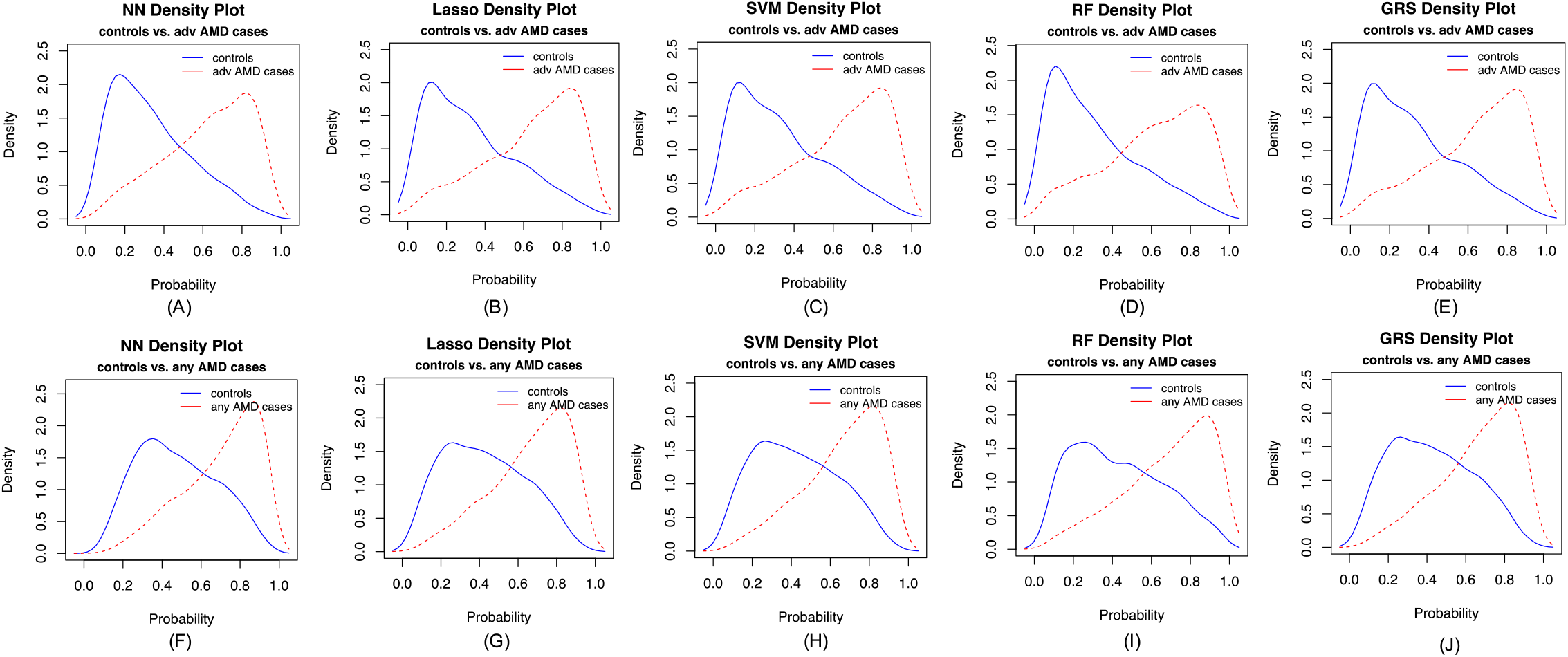
Density curves of the predicted risk for the two counterparts for five prediction methods. (A-E) normal controls vs. advanced AMD cases, and (F-J) normal controls vs. any AMD cases.

**Figure 4.**
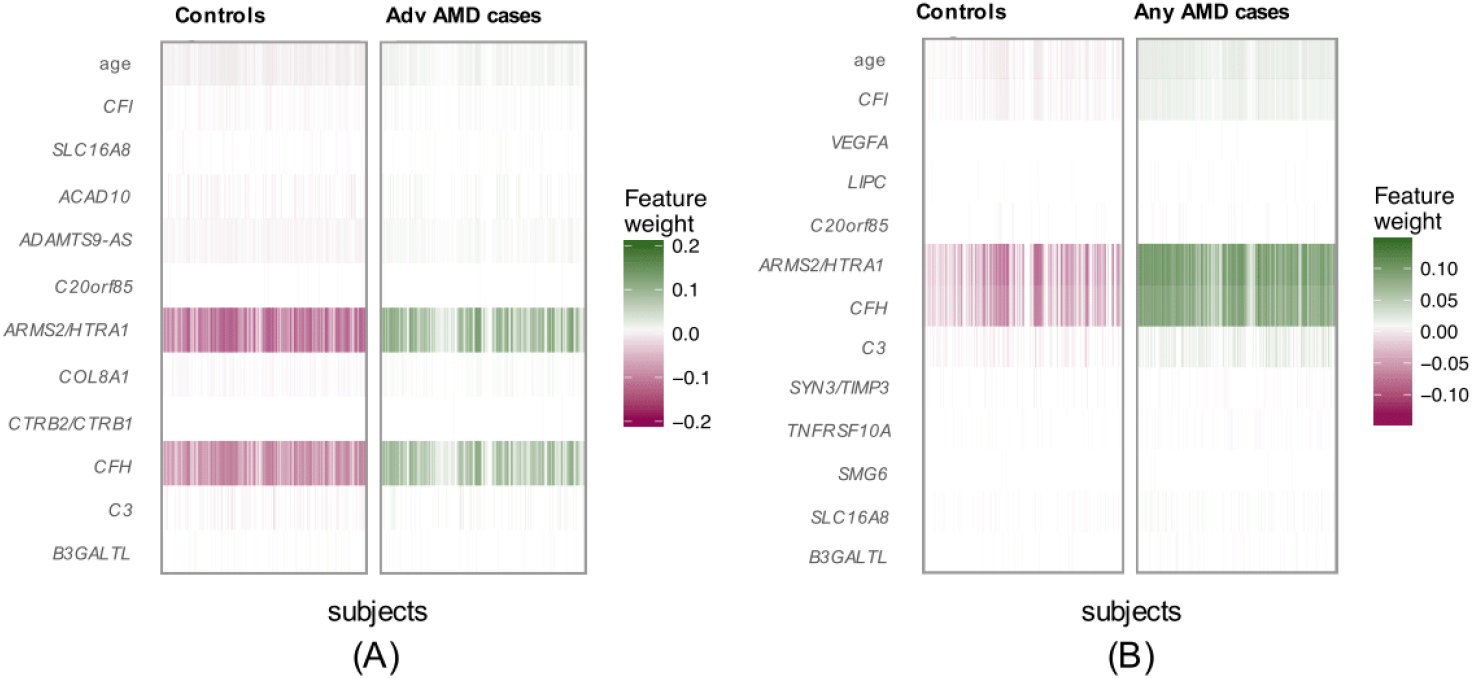
Feature importance heatmaps from LIME for NN. (A) Scenario 1: normal controls vs. advanced AMD cases, and (B) Scenario 2: normal controls vs. any AMD cases.

In the secondary list of feature SNPs, we used all genome-wide significant loci and age as predictors. We applied NN and Lasso, but SVM and RF were excluded because they are not suitable for a large number of predictors. GRS was also excluded, because a large group of less significant SNPs in linkage disequilibrium (LD) may contribute more to the prediction than a single very significant SNP. On the contrary, NN and Lasso assigned penalties to the highly correlated features, which adjusted for the correlations among SNPs in high LD. Although the results were similar to the previous parsimonious models, NN showed slightly better AUCs than Lasso by an average of 0.01 (Figure S8 and Table S2). In another set of secondary list of feature SNPs with *P* < 1×10^−5^, the prediction accuracy did not improve in terms of AUCs (Figure S10). In addition, we evaluated the performance in a non-Caucasian test dataset from the same dbGaP project to assess whether our training results from Caucasians could be applied to non-Caucasians. This non-Caucasian test dataset included a mixed population of Africans, Asians and unknown ancestry subjects. The results (Table S3) showed that the prediction is obviously worse in non-Caucasians (e.g., AUC = 0.72∼0.74 in NN) than in Caucasians (e.g., AUC = 0.82∼0.83 in NN).

All five prediction approaches provided similar results. The prediction between normal controls and advanced AMD had good performance. However, the prediction between normal controls and any AMD could be more useful for covering subjects with all possible AMD statuses in the training process. In addition, the parsimonious models with only one top SNP from each significant locus could be easier to conduct in practice than including all significant SNPs.

In addition to the test dataset we generated from dbGaP, we extracted a set of 383 subjects with macular degeneration (i.e., all cases) and above 50 years from the UK Biobank^9^ as an independent test dataset. Since all approaches seem to give similar results, we used NN to evaluate the model performance in this independent cohort. It is not surprising that the accuracy equal to 0.46 (i.e., sensitivity for this test set with all samples being AMD cases) was not good, because their age (age = 62.30 ± 5.61) was much younger than the training dataset (age = 76.32 ± 8.24). Since AMD is irreversible, these UK Biobank subjects will still be AMD cases in their future years. Thus, we added 14 years to each subject to match the age range in our training dataset, and the accuracy increased to 0.77. If we excluded age from the model and only kept SNPs, the accuracy was equal to 0.75, which is still quite acceptable.

## DISCUSSION

AMD is one of the most successful diseases for GWAS with multiple consistently replicated loci. The dbGaP (phs001039.v1.p1) dataset from the International Age-Related Macular Degeneration Genomics Consortium is the largest publicly available genotype dataset by far with 35,358 subjects. Our prediction results show that SNPs along with age could predict AMD status with very good accuracy in Caucasians.

We did not directly use the 52 SNPs from 34 reported loci from Fritsch *et al*. ^5^ as predictors, since use of these loci could lead to over-fitting, as they were identified using all data including both training and test data. To select our feature SNPs for predictions, we conducted GWAS for four scenarios comparing among normal controls, intermediate AMD and advanced AMD cases. To our knowledge, these are the first large GWAS accounting for intermediate AMD. Most of the genome-wide significant SNPs from these four GWAS were identified from the previous large AMD GWAS ^5^. However, *ABHD2* from chromosome 15 was identified for the first time in the comparison between intermediate AMD and advanced AMD; and *TNFRSF10A* from chromosome 8 and *SMG6* from chromosome 17 were identified in the comparison between normal controls and any AMD cases. They were not observed previously, because only the comparison between no AMD and advanced AMD was studied before. These AMD related genome-wide significant SNPs (*P*<5×10^−8^) were used for predictions. We observed that including more SNPs (*P*<1×10^−5^) did not improve the prediction performance in the test set, because more features could lead to overfitting in the training set. We also conducted a fifth scenario comparing dry and wet AMD cases (results not shown). The SNPs from *ARMS2/HTRA1* and *MMP9* showed significantly different genetic effects between dry and wet AMD. However, these two genes were not able to classify dry and wet AMD. For the reference, we also used the 52 reported SNPs ^5^ in our prediction models and the results showed slightly better prediction accuracy than our selected feature SNPs (Figure S11). This could be due to the use of test data information in the training step.

It is not surprising that the comparison between normal controls and advanced AMD detected the most significant genetic loci, which was also the comparison used in the recent large AMD GWAS ^5^, and this comparison also achieved the highest prediction accuracy, because the difference of clinical characteristics between no and advanced AMD is more obvious than comparing them to intermediate AMD. Power could be another reason due to a smaller sample size of intermediate AMD than no and advanced AMD. However, the comparison between normal controls and any AMD could be more clinically useful than the comparison between normal controls and advanced AMD, because the training process covers all possible outcomes (i.e., no, intermediate and advanced AMD) so as to be able to predict any subject.

In this study, we considered five prediction methods: NN, Lasso, SVM, RF and GRS. In the primary list of feature SNPs, they all achieved similar prediction accuracy. In the secondary list of feature SNPs (Figures S8 and S10), NN consistently had slightly higher AUCs than Lasso. One of the advantages that NN has as compared to Lasso is that NN accounts for nonlinear relationship and interactions among predictors in addition to linear relationship. On the contrary, Lasso only accounts for linear relationship. The NN is equivalent to Lasso when only input and output layers are included with L1 norm at the input layer (Figure S1).

Age is an important predictor. A logistic regression showed that age alone could achieve moderate accuracy for predicting AMD risk (Figure S12). Moreover, since age is shown to be an important predictor, the predictive models established from all five approaches could predict a person’s AMD risk at any future age above 50 years.

Our study has some limitations. It could be more useful to predict the time to AMD progression instead of predicting the AMD risk, since the AMD status may change if the follow-up time is extended. In this study, NN does not show a great advantage over other competing approaches, which suggests that most of prediction approaches could achieve similar results for predicting AMD risk with the same set of predictors. NN might be more advantageous than other approaches when using large number of input features because it is considered to be flexible and can handle large sets of predictors (given enough data). We have implemented the established prediction model from the NN approach for normal controls versus any AMD using R Shiny, which is available at https://yanq.shinyapps.io/no_vs_amd_NN/. Note that the final predicted AMD risk output from this app is adjusted for population prevalence (see supplementary text for details).

## Data Availability

The study subjects are from the International Age-Related Macular Degeneration Genomics Consortium - Exome Chip Experiment dbGaP data set (phs001039.v1.p1).

https://www.ncbi.nlm.nih.gov/projects/gap/cgi-bin/study.cgi?study_id=phs001039.v1.p1

## Notes

### Competing Interest Statement

The authors have declared no competing interest.

### Funding Statement

No external funding was received.

### Author Declarations

All relevant ethical guidelines have been followed and any necessary IRB and/or ethics committee approvals have been obtained.

Any clinical trials involved have been registered with an ICMJE-approved registry such as ClinicalTrials.gov and the trial ID is included in the manuscript.

